# Detecting cognitive decline using speech only: The ADReSS_*O*_ Challenge

**DOI:** 10.1101/2021.03.24.21254263

**Authors:** Saturnino Luz, Fasih Haider, Sofia de la Fuente, Davida Fromm, Brian MacWhinney

## Abstract

Building on the success of the ADReSS Challenge at Interspeech 2020, which attracted the participation of 34 teams from across the world, the ADReSS_*o*_ Challenge targets three difficult automatic prediction problems of societal and medical relevance, namely: detection of Alzheimer’s Dementia, inference of cognitive testing scores, and prediction of cognitive decline. This paper presents these prediction tasks in detail, describes the datasets used, and reports the results of the baseline classification and regression models we developed for each task. A combination of acoustic and linguistic features extracted directly from audio recordings, without human intervention, yielded a baseline accuracy of 78.87% for the AD classification task, a root mean squared (RMSE) error of 5.28 for prediction of cognitive scores, and 68.75% accuracy for the cognitive decline prediction task.

## 1. Introduction

Dementia is a category of neurodegenerative diseases which entail long-term and usually gradual decrease of cognitive functioning. The main risk factor for dementia is age and, hence, it is increasingly prevalent in our ageing society. Due to the severity of the disease, institutions and researchers worldwide are investing considerably on dementia prevention, early detection and disease progression monitoring [1]. There is a need for cost-effective and scalable methods for detection of early signs of Alzheimer’s Dementia (AD) as well as prediction of disease progression.

Methods for screening and tracking the progression of dementia traditionally involve cognitive tests such as the MiniMental Status Examination (MMSE) [2] and the Montreal Cognitive Assessment (MoCA) [3]. MMSE and MoCA are widely used because, unlike imaging methods, they are cheap, quick to administer and easy to score. Despite its shortcomings in specificity in early stages of dementia, the MMSE is still widely used [4]. The promise of speech technology in comparison to cognitive tests is twofold. First, speech can be collected passively, naturally and continuously throughout the day, gathering increasing data points while burdening neither the participant nor the researcher. Furthermore, the combination of speech technology and machine learning creates opportunities for automatic screening and diagnosis support systems for dementia. The ADReSS_*o*_ Challenge aims to generate systematic evidence for these promises towards their clinical implementation.

As with its predecessor, the overall objective of the ADReSS_*o*_ Challenge is to host a shared task for the systematic comparison of approaches to the detection of cognitive impairment and decline based on spontaneous speech. As has been pointed out elsewhere [5, 6], the lack of common, standardised datasets and tasks has hindered the benchmarking of the various approaches proposed to date, resulting in a lack of translation of these speech based methods into clinical practice.

The ADReSS_*o*_ Challenge provides a forum for researchers working on approaches to cognitive decline detection based on speech data to test their existing methods or develop novel approaches on a new shared standardised dataset. The approaches that performed best on last year’s dataset [5] employed features extracted from manual transcripts which were provided along with the audio data [7, 8]. The best performing method [8] made interesting use of pause and disfluency annotation provided with the transcripts. While this provided interesting insights into the predictive power of these paralinguistic features for detection of cognitive decline, extracting such features, and indeed accurate transcripts from spontaneous speech remains an open research issue. This year’s ADReSS_*o*_ (Alzheimer’s Dementia Recognition through Spontaneous Speech *only*) tasks provide more challenging and improved spontaneous speech datasets, requiring the creation of models straight from speech, without manual transcription, though automatic transcription is encouraged.

The ADReSS_*o*_ datasets are carefully matched so as to avoid common biases often overlooked in evaluations of AD detection methods, including repeated occurrences of speech from the same participant (common in longitudinal datasets), variations in audio quality, and imbalances of gender and age distribution. The challenge defines three tasks:

1. an AD classification task, where participants were required to produce a model to predict the label (AD or non-AD) for a short speech session. Participants could use the speech signal directly (acoustic features), or attempt to convert the speech into text automatically (ASR) and extract linguistic features from this automatically generated transcript;
2. an MMSE score regression task, where participants were asked to create models to infer the patients’ MMSE score based solely on speech data; and
3. a cognitive decline (disease progression) inference task, where they created models for prediction of changes in cognitive status over time, for a given speaker, based on speech data collected at baseline (i.e. the beginning of a cohort study).

These tasks depart from neuropsychological and clinical evaluation approaches that have employed speech and language [9] by focusing on prediction and recognition using spontaneous speech. Spontaneous speech analysis has the potential to enable novel applications for speech technology in longitudinal, unobtrusive monitoring of cognitive health [10], in line with the theme of this year’s INTERSPEECH, “Speech Everywhere!”.

This paper describes the ADReSS_*o*_ dataset and presents baselines for all ADReSS_*o*_ tasks, including feature extraction procedures and models for AD detection, MMSE score regression and prediction of cognitive decline.

## 2. Related work

There has been increasing research on speech technology for dementia detection over the last decade. The majority of this research has focused on AD classification, but some of it targets MCI detection as well [6]. These objectives are most closely related with our first task, namely, the AD classification task. Such related research includes the best performing models presented in the ADReSS challenge in 2020. These achieved an 85.45% [7] and 89.6% [8] accuracy in AD classification using acoustic features and text-based features extracted from manual transcripts. Classification based on acoustic features only was also attempted in [7], and obtained 76.85% accuracy with the IS10-Paralinguistics feature set (a low dimensional version of ComParE [11]) and Bag-of-Acoustic-Words (BoAW).

Few works rely exclusively on acoustic features or text features extracted through ASR. One of these achieved a 78.7% accuracy on a subset of the Cookie Theft task of the Pitt dataset, using different comprehensive paralinguistic feature sets and standard machine learning algorithms [12]. Another, using the complete Pitt dataset, obtained 68% accuracy using only vocalisation features (i.e. speech-silence patterns) [10]. A classification accuracy of 62.3% was reported in a study that used fully automated ASR features with a different dataset [13].

As regards the second task, regression over MMSE scores, there is less literature available and most of it has been presented in recent workshops [6]. Several of these works used the above mentioned Pitt dataset to extract different linguistic and acoustic features and predict MMSE scores. A recent study captured different levels of cognitive impairment with a multiview embedding and obtained a mean absolute error (MAE) of 3.42 [14]. Another study reported a MAE of 3.1, having relied solely on acoustic features to build their regression model (a set of 811 features) [15]. Error scores as low as 2.2 (MAE) have been obtained, but relying on non-spontaneous speech data elicited in semantic verbal fluency (SVF) tasks [16].

Studies addressing the progression task are far less common. Notable in this category is [17], which incorporated a comprehensive set of features (i.e. lexicosyntactic, semantic and acoustic) into Bayesian network with, reporting a MAE of 3.83 on prediction of MMSE scores throughout different study visits. Two other studies account for disease progression in classification experiments. One of them extracted speech-based from the ISLE dataset achieving 80.4% accuracy to detect intra-subject cognitive changes, that is, to distinguish healthy participants who remained healthy from those who developed some kind of cognitive impairment [18]. The second study uses SVF scores to build a machine learning classifier able to predict changes from MCI to AD over a 4-year follow-up, with 84.1% accuracy [19].

## 3. The ADReSS_*o*_ Datasets

We provided two distinct datasets for the ADReSS_*o*_ Challenge:

1. a dataset consisting of speech recordings of Alzheimer’s patients performing a category (semantic) fluency task [20] at their baseline visit, for prediction of cognitive decline over a two year period, and
2. a set of recordings of picture descriptions produced by cognitively normal subjects and patients with an AD diagnosis, who were asked to describe the Cookie Theft picture from the Boston Diagnostic Aphasia Examination [21, 22].

The recorded data also included speech from different experimenters who gave instructions to the patients and occasionally interacted with them in short dialogues. No transcripts were provided with either dataset, but segmentation of the recordings into vocalisation sequences with speaker identifiers [23] were made available for optional use. The ADReSS_*o*_ challenge’s participants were asked to specify whether they made use of these segmentation profiles in their predictive modelling. Recordings were acoustically enhanced with stationary noise removal and audio volume normalisation was applied across all speech segments to control for variation caused by recording conditions such as microphone placement.

The dataset used for AD and MMSE prediction was matched for age and gender so as to minimise risk of bias in the prediction tasks. We matched the data using a propensity score approach [24, 25] implemented in the R package MatchIt [26]. The final dataset matched according to propensity scores defined in terms of the probability of an instance of being treated as AD given covariates age and gender. All standardised mean differences for the age and gender covariates were *<* 0.001 and all standardised mean differences for age^2^ and two-way interactions between covariates were well below .1, indicating adequate balance for the covariates. The propensity score was estimated using a probit regression of the treatment on the covariates age and gender as probit generated a better balanced than logistic regression. The age/gender matching is summarised in Figure 1, which shows the respective (empirical) quantile-quantile plots for the original and balanced datasets. A quantile-quantile plot showing instances near the diagonal indicates good balance.

**Figure 1:**
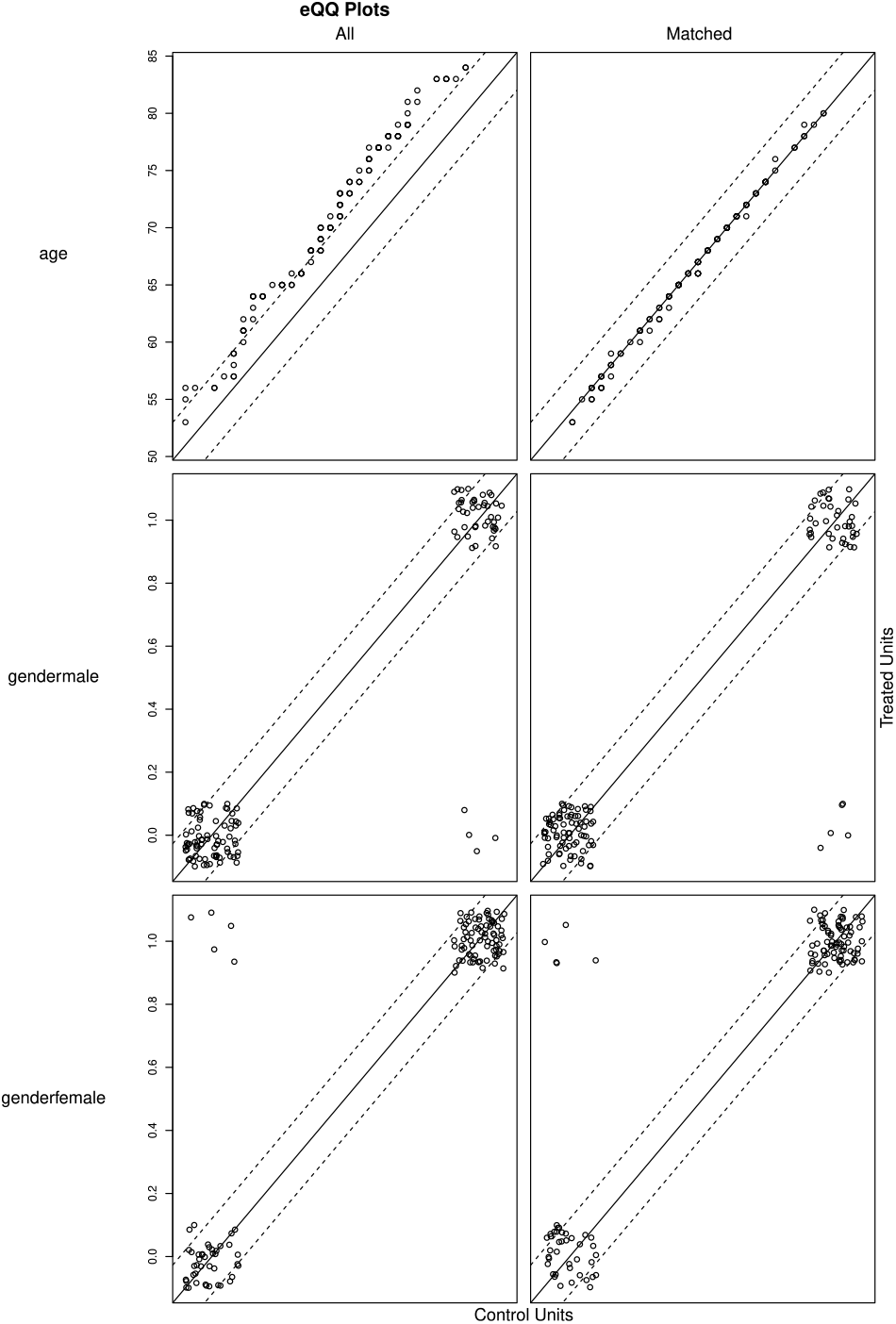
Quantile-quantile plots for data before (left) and after matching (right) by age and gender.

The resulting dataset encompasses 237 audio files. These were split into training and test sets, with 70% of instances allocated to the former and 30% allocated to the latter. These partitions were generated so as to preserve gender and age matching.

The dataset for the disease prognostics task (prediction of cognitive decline) was created from a longitudinal cohort study involving AD patients. The time period for assessment of disease progression spanned the baseline and the year-2 data collection visits of the patients to the clinic. The task involves classifying patients into ‘decline’ or ‘no-decline’ categories, given speech collected at baseline as part of a verbal fluency test. Decline was defined as a difference in MMSE score between baseline and year-2 greater than or equal 5 points (i.e. *mmse*(*baseline*) *— mmse*(*y*2) *≥* 5). This dataset has a total of 105 audio recordings split into training and test sets as with the diagnosis dataset (70% and 30% of recordings, respectively).

## 4. Data representation

### 4.1. Acoustic features

We applied a sliding window with a length of 100 ms on the audio files of the dataset with no overlap and extracted *eGeMAPS* features over such frames. The *eGeMAPS* feature set [27] resulted from an attempt to reduce the somewhat unwieldy feature sets above to a basic set of acoustic features based on their potential to detect physiological changes in voice production, as well as theoretical significance and proven usefulness in previous studies. It contains the F0 semitone, loudness, spectral flux, MFCC, jitter, shimmer, F1, F2, F3, alpha ratio, Hammarberg index and slope V0 features, as well as their most common statistical functionals, totalling 88 features per 100ms frame. We then applied the active data representation method (ADR) [12] to generate a data representation using frame level acoustic information for each audio recording. The ADR method has been tested previously for generating representations for large scale time-series data. It employs self-organising mapping to cluster the original acoustic features and then computes second-order features over these cluster to extract new features (see [12] for details). Note that this method is entirely automatic in that no speech segmentation or diarisation information is provided to the algorithm.

### 4.2. Linguistic Features

We used the Google Cloud-based Speech Recogniser for automatically transcribing the audio files. The transcripts were converted into CHAT format which is compatible with CLAN [28], a set of programs that allows for automatic analysis of a wide range of linguistic and discourse structures. Next, we used the automated MOR function to assign lexical and morphological descriptions to all the words in the transcripts. Then, we used two commands: EVAL which creates a composite profile of 34 measures, and FREQ to compute the Moving Average Type Token Ratio [29].

## 5. Diagnosis baseline

### 5.1. Task 1: AD Classification

The AD classification experiments were performed using five different methods, namely decision trees (DT, where the leaf size is optimised through a grid search within a range of 1 to 20), nearest neighbour (KNN, where K parameter is optimised through a grid search within a range of 1 to 20), linear discriminant analysis (LDA), Tree Bagger (TB, with 50 trees, where leaf size is optimised through a grid search within a range of 1 to 20), and support vector machines (SVM, with a linear kernel, where box constraint is optimised by trying a grid search between 0.1 to 1.0, and a sequential minimal optimisation solver).

The results for accuracy in the AD vs Control (CN) classification task are summarised in Table 1. As indicated in bold-face, the best performing classifier in cross-validation (CV) was DT, achieving 78.92% and 72.89% accuracy using acoustic and linguistic features, respectively. On the test set, however, the results were reversed, with linguistic features producing an overall best accuracy of 77.46%, with the SVM classifier. Late fusion of the acoustic and linguistic models improves the accuracy on the test set further to 78.87% (Figure 2).

**Table 1:** Task1: AD classification accuracy on leave-one-subject out CV and test data.

|  |  | LDA | DT | SVM | TB | KNN | mean (sd) |
| --- | --- | --- | --- | --- | --- | --- | --- |
| CV | Acoustic | 62.65 | **78.92** | 69.28 | 65.06 | 65.06 | 68.19 (6.4) |
|  | Linguistic | 72.29 | **72.89** | 72.89 | 75.90 | 65.06 | 71.81 (4.0) |
| Test | Acoustic | 50.70 | 60.56 | **64.79** | 63.38 | 53.52 | 58.59 (6.2) |
|  | Linguistic | 76.06 | 74.65 | **77.46** | 73.24 | 59.15 | 72.11 (7.4) |

**Figure 2:**
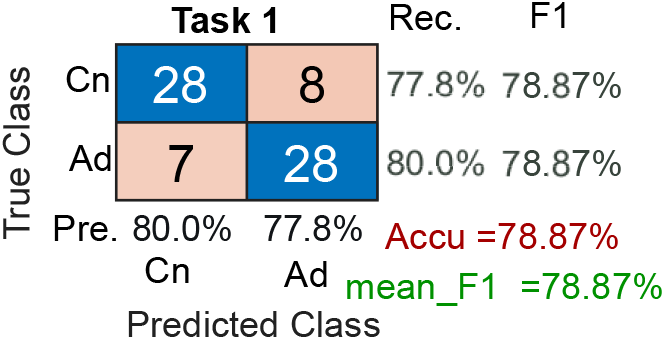
Task 1: Late (decision) fusion of the best results of acoustic and linguistic models. Precision (Pre), recall (Rec), accuracy (Accu) and mean F_1_ scores are shown on the margins.

### 5.2. Task 2: MMSE prediction

For this regression task we also used five types of regression models: linear regression (LR), DT, with leaf size of 20 and CART algorithm, support vector regression (SVR, with a radial basis function kernel with box constraint of 0.1, and sequential minimal optimisation solver), Random Forest regression ensembles (RF), and Gaussian process regression (GP, with a squared exponential kernel). The regression methods are implemented in MATLAB [30] using the statistics and machine learning toolbox.

The results are summarised in Table 2. As with classification, DT regression outperformed the other models in CV, with ASR linguistic features outperforming acoustic ADR features. This trend persisted in the test set, with linguistic features producing a minimum RMSE of 5.28 in a SVR model. We then fused the best results of linguistic and acoustic features and took a weighted mean, finding the weights through grid search on the validation results, which resulted in an improvement (6.37) on the validation dataset. We then used the same weights to fuse the test results and obtained an RMSE of 5.29 (*r* = 0.69).

**Table 2:** Task2: MMSE score prediction error scores (RMSE) for leave-one-subject out CV and test data.

|  | LR | DT | SVR | RF | GP | mean (sd) |
| --- | --- | --- | --- | --- | --- | --- |
| CV Acoustic | 6.88 | 6.88 | 6.96 | 7.89 | 6.71 | 7.06 (0.47) |
| CV Linguistic | 6.65 | 5.92 | 6.42 | 7.02 | 6.50 | 6.50 (0.40) |
| Test Acoustic | 6.23 | 6.47 | **6.09** | 8.18 | 6.81 | 6.75 (0.84) |
| Test Linguistic | 5.87 | 6.24 | **5.28** | 6.94 | 5.43 | 5.95 (0.67) |

## 6. Prognosis baseline

### 6.1. Task 3: prediction of progression

We tested the same classification methods used in Task 1 for the task of identifying those patients who went on to exhibit cognitive decline within two years of the baseline visit in which the speech samples used in our models were taken. The acoustic and linguistic features were generated as described in Section 4. The results of this prediction task are summarised in Table 3. As the classes for this task are imbalanced we report average *F*_1_ results rather than accuracy, Once again DT performed best on CV, but the *F*_1_ results for the test set was considerably lower, reaching only a maximum of 66.67%, for linguistic features and 61.02% for acoustic features.

**Table 3:** Task3: cognitive decline progression results (mean of F_1_Score) for leave-one-subject-out CV and test data.

|  | LDA | DT | SVM | TB | KNN | mean (sd) |
| --- | --- | --- | --- | --- | --- | --- |
| Val Acoustic | 59.89 | 84.94 | 55.64 | 63.85 | 65.92 | 66.05 (11.27) |
| Val Linguistic | 55.19 | 76.52 | 45.24 | 63.10 | 55.25 | 59.06 (11.64) |
| test Acoustic | **61.02** | 53.62 | 40.74 | 40.74 | 38.46 | 46.91 (9.89) |
| test Linguistic | 54.29 | **66.67** | 40.74 | 56.56 | 39.62 | 51.58 (11.41) |

As before, we fused the predictions of the best models for each feature type, hoping that the diversity of models might improve classification. The confusion matrix for the fusion model is shown in Figure 3. This time, however, decision fusion did not yield any improvement in accuracy. However, it is noted that the recall (sensitivity) improved from 40% to 70% for patients exhibiting cognitive decline.

**Figure 3:**
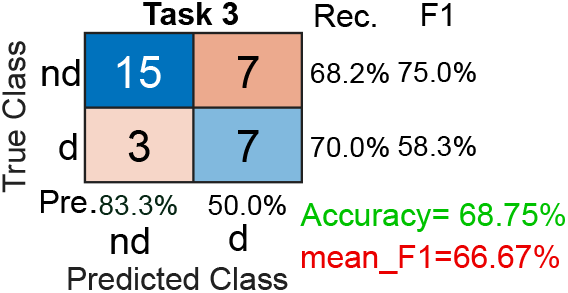
Task 3: Decision fusion of the best results of acoustic and linguistic features on the test set.

## 7. Discussion

The AD classification baseline yielded a maximum accuracy of 78.87% on the test set, through the fusion of models based on linguistic and acoustic features. Despite the fact that the ASR transcripts had relatively high word error rates, linguistic features contributed considerably to the predictions. The overall baseline results for this task are in fact comparable to results obtained for similar picture description data using manual transcripts (see Section 2). DT classifiers performed well on the CV experiments, but accuracy decreased on the test set, indicating probable overfitting. Overall, however, all models proved fairly robust.

A similar picture was observed in the MMSE regression task. Linguistic features contributed appreciably to the prediction, even though the transcripts contained many errors. In this case, however, late fusion only improved the RMSE score in CV; the test set RMSE remained practically unchanged.

The prognosis task proved to be the most difficult prediction task. The CV results varied considerably among models, with a standard deviation of 11.64 for the linguistic models. The test set results were also varied, reaching a maximum *F*_1_ score of 66.67%, even when the best model predictions were fused. Although the acoustic features produced the best classification results in CV (*F*_1_ = 66.05% vs 59.06% for linguistic features), these results were not born out by test set evaluation, suggesting that the acoustic features made the classifiers more prone to overfitting. It is possible that this could be mitigated by training the acoustic feature extractor (ADR) on a larger set of off-task recordings (data augmentation) and fine tuning the resulting model on the ADReSS_*o*_ data.

## 8. Conclusions

The ADReSS_*o*_ Challenge is the first shared task to target cognitive status prediction using raw, non-annotated a non-transcribed speech, and to address prediction of changes in cognition over time. We believe this moves the speech processing and machine learning methods one step closer to the real-world of clinical applications. A limitation the AD classification and the MMSE regression tasks share with most approaches to the use of these methods in dementia research is that they provide little insight into disease progression. This has been identified as the main issue hindering translation of these technologies into clinical practice [6] and, hence, preclinical modelling emerges as clear avenue for future research [31]. However, these tasks remain relevant in application scenarios involving automatic cognitive status monitoring, in combination with wearable and ambient technology. The addition of the progression task should open avenues for relevance also in more traditional clinical contexts.

## Data Availability

All data used in this paper has been made available through DementiaBank, as part of the ADReSSo challenge.

https://dementia.talkbank.org/

## 9. Acknowledgements

This research is funded by the European Union’s Horizon 2020 research programme, under grant agreement 769661, SAAM project. SdlFG is supported by the Medical Research Council.

